# Associations between patient characteristics at referral and social worker care coordinator allocation in mental healthcare: a retrospective cohort study

**DOI:** 10.64898/2026.10.29.26366330

**Authors:** Norah Alothman, Matthew Broadbent, Gayan Perera, Robert Stewart

## Abstract

**Objectives:** To investigate care coordination delivered specifically by social workers and, more generally, by other professionals using electronic health records.

**Study design and Setting:** A retrospective cohort study in a large mental healthcare provider serving a South London catchment.

**Sample:** The study comprised care coordination episodes from 2007-2023 for adults aged 18-65 years at the time of commencement. We categorised these by the care coordinator’s profession: social worker, nurse, doctor, and others, with a focus in the analysis on social worker care coordination. In addition to sociodemographic data, we obtained diagnosis, mental health service use, and symptoms over the 12 months preceding episode start.

**Outcome measures:** Social workers’ associations with mental healthcare service use 12 months before the episode start. Cases allocated to social workers the year after had higher rates across most service-use outcomes.

**Results:** Of 327,056 care coordination episodes for 132,553 patients, 10.6% had a social worker care coordinator. Full-adjusted associations with social worker care coordinator included longer duration since first referral (IRR 1.05, 95% CI 1.03-1.07, P <0.001), more prior hospitalisations involving compulsory detention (1.11, 1.07-1.15, P <0.001), more days under a home treatment team (1.25, 1.17-1.34, P <0.001), and more emergency department mental health contacts (1.10, 1.07-1.14, P <0.001), as well as higher recorded symptoms on all scales.

**Conclusion:** Episodes with social workers as care coordinators are associated with more complex needs, severe symptoms and a later stage in the course of care. The assignment of care coordinators by profession and its influence on clinical outcomes remain unclear; further mixed-methods research could help clarify this.

**Article Summary:** *Strengths and limitations of this study:*

- We believe this is the first large quantitative study evaluating the care coordination delivered by social workers specifically or between different professional groups.
- The study incorporates an evaluation of symptom profiles utilising natural language processing.
- Our study utilised a large sample from routine mental healthcare over a long period from a highly socially and culturally diverse catchment.
- This study from a single catchment and provider.
- Comparisons relied on the assigning of a profession to each care coordinator.

## Background

In England, more than 3 million people had contact with secondary mental health, autism or learning disability services in 2022/23, 24% higher than in 2019/20.^1^ People with mental disorders often require structured support, as they are most likely to need access to a range of services inside or outside the healthcare provider.^2–4^ A recent report from the House of Commons Committee for Adult Social Care Reform highlighted that there are 1.5 million people of working age in England not receiving the care they need, which impacts the quality of their lives.^5^ Moreover, studies have shown that people often report challenges in accessing the necessary services, which may negatively impact on their health outcomes.^4,5^ Therefore, care coordinators are fundamental to high-functioning healthcare services more generally and have long been core to the services that are provided in mental healthcare.^3,6^

Care coordination comprises organised interactions and activities between different service providers that include appropriate time transfer and precise information with effective communication.^3,6–8^ Studies have defined care coordination as the planned organisation of patient care activities among different professionals to ensure that patients receive appropriate care that meets their needs.^3,6,9^ The care coordination role is particularly important for patients requiring longer treatment episodes.^3,10^ While care coordination interventions can take different forms, the primary aim is typically to facilitate communication and case management.^3^ Traditionally, the role of the care coordinator is provided by health or social care professionals, including nurses, doctors and occupational therapists, but also social workers, as this is historically part of social work practice.^7,10,11^ Additionally, alongside the treatment of the illness, social workers employed in mental healthcare services are particularly qualified to work with people on issues such as housing, occupation, and sources of income, which usually requires coordination between different services.^4^ Thus, social work care coordination can play an essential role, focusing on patients’ needs and helping them access needed services promptly to avoid health crises.^4,7,10^

The role of the care coordinator is central to community mental health service operation. However, there has been little quantitative evaluation of care coordination delivered by social workers specifically or between different professional groups more generally. As the beginning of a wider programme of research, we sought to generate evidence on this using large routine mental healthcare data, initially investigating associations between patient characteristics and care coordinator allocations by profession.

## Methods

### Setting and data sources

Data were assembled from the South London and Maudsley NHS Foundation Trust (SLaM): one of Europe’s largest mental healthcare providers, covering four South London boroughs (Croydon, Lambeth, Lewisham, and Southwark) with around 1.3 million residents. SLaM has over 240 services with around 6000 staff for this catchment in addition to substance misuse services for people living in a wider geography.^12^ The Trust has used electronic health records since 2006, and the Clinical Record Interactive Search (CRIS) was set up as a platform to provide researcher access to deidentified data from the source records. CRIS allows researchers to assemble and conduct secondary analyses using data from over 500,000 cases since 2006, supplemented by extensive meta-data derived from natural language processing (NLP) algorithms applied to unstructured text fields, and by a range of linkages to external data resources. CRIS has received full ethical approval as an anonymised data resource (Oxford Research Ethics Committee C, reference 23/SC/0257).^13^ This study was conducted under the CRIS database approval. Therefore, individual informed consent was not required, as the national data governance law permits and governs the use of anonymised health care data for research purposes without specific consent.

### Sample

The study sample comprised care coordination episodes commencing between 1st January 2007 and 31st December 2023, and these episodes were categorised according to the profession of the allocated care coordinator under the following groups: social worker, nurse, medical doctor, and others. These groups were ascertained from the ‘professional Group ID’ on the system associated with the generic documents assembled for the patient records.^14^ The sample was restricted to patients aged 18 to 65 years old at the start date of the episode, and this start date was defined as the index date for covariate definitions.

### Measurements

From extracted data, demographic and clinical characteristics were defined from measures at or closest to the index date, with service use and mental health symptom scores calculated from available records over the 12 months prior to the index date. Sociodemographic characteristics comprised age, gender, marital status, ethnicity, cohabitation status (living alone or not, derived via an NLP algorithm), and neighbourhood deprivation quantified by the Index of Multiple Deprivation (IMD), a variable derived from multiple domains collected in the national census and applied at the level of the Lower Super Output Area, a standard administrative unit containing around 1500 residents. Primary mental health diagnosis was also ascertained, coded according to the International Classification of Diseases, 10^th^ edition (ICD-10). Health of the Nation Outcome Scales (HoNOS) were extracted based on the HoNOS recorded closest to the index date; these are routine scales in widespread use assessing the health and social performance of people with mental disorders and scores on a 0-4 scale were categorised into binary variables (0-1 no/borderline vs. 2-4 mild-severe problems). We also obtained information on the duration of time since first referral to SLaM, the subsequent duration of the index care coordination episode and the following service use measures over the 12 months preceding the index date: i) number of acute admissions and ii) those involving a Mental Health Act section, iii) days under a home treatment team, iv) days in mental health inpatient care, v) number of face-to-face community contacts and vi) number of face-to-face mental health service emergency department contacts. Finally, using over 60 NLP algorithms, we ascertained the number of different symptoms recorded at any point in the 12 months prior to the index date and scaled these within the following categories: positive psychotic, negative psychotic, depressive, manic, disorganised, and catatonic.

### Statistical analyses

Statistical analyses were carried out using STATA version 17. The unit of the analysis was the care coordination episode; episodes with missing professional group IDs were excluded from the analyses. Initial descriptive analyses were used to summarise the distribution of demographic and clinical characteristics. Chi-squared tests were used to compare age group, gender, marital status, ethnicity, deprivation quintile, primary diagnosis and HoNOS subscale binary variables – initially for heterogeneity across the care coordinator professional groups and then for primary analyses between social worker and other care coordinators. ANOVA and t-tests were used for the same two comparisons for remaining continuously distributed variables. To take into account the multiple episodes occurring in the same individual, clustering was applied using Generalised Estimating Equations (GEE) with a negative binomial regression model, as the count outcomes were overdispersed. An exchangeable correlation was used, and for models that failed to converge, an independent structure correlation with Huber-White cluster-robust sandwich standard errors was used. Analyses were then conducted to calculate the incidence rate ratios (IRRs) and p-values for the service use and symptom scale measures as count dependent variables, comparing social worker vs. other care coordinators. Unadjusted regression models were followed by those adjusted for gender, ethnicity, marital status, age, IMD and for primary diagnosis, and then by symptom scales for service use outcomes, or by service use measures for symptom scales as outcomes. Logistic regression models were applied for the HoNOS scores as binary variable to report the odds ratios (ORs) with 95% confidence intervals (Cls) and p-values.

### Patient and public involvement

At the project planning stage, members of a Service User Advisory Group were shown initial ideas and plans and asked for thoughts and suggestions relating to a broader body of work (a PhD programme for NA) of which this study was a component. The group consisted of service users (of SLaM services) with lived experience of mental health problems. They were very keen on finding out the results of the first author’s PhD study. Suggestions were taken on board and informed, in part, some of the research questions.

## Results

A total 327,056 recorded care coordination episodes were ascertained for 132,553 patients over a 17-year period. Table 1 describes characteristics according to the profession of the care coordinator. All characteristics varied significantly across the four professional groups. For episodes involving social worker care coordination, patients were more likely to be in older age groups, male, non-cohabiting, from non-White ethnic groups, and living in a higher deprivation neighbourhood. Primary diagnoses for these episodes were more likely to be schizophrenia-related, mood disorders, or unspecified.

**Table 1.**
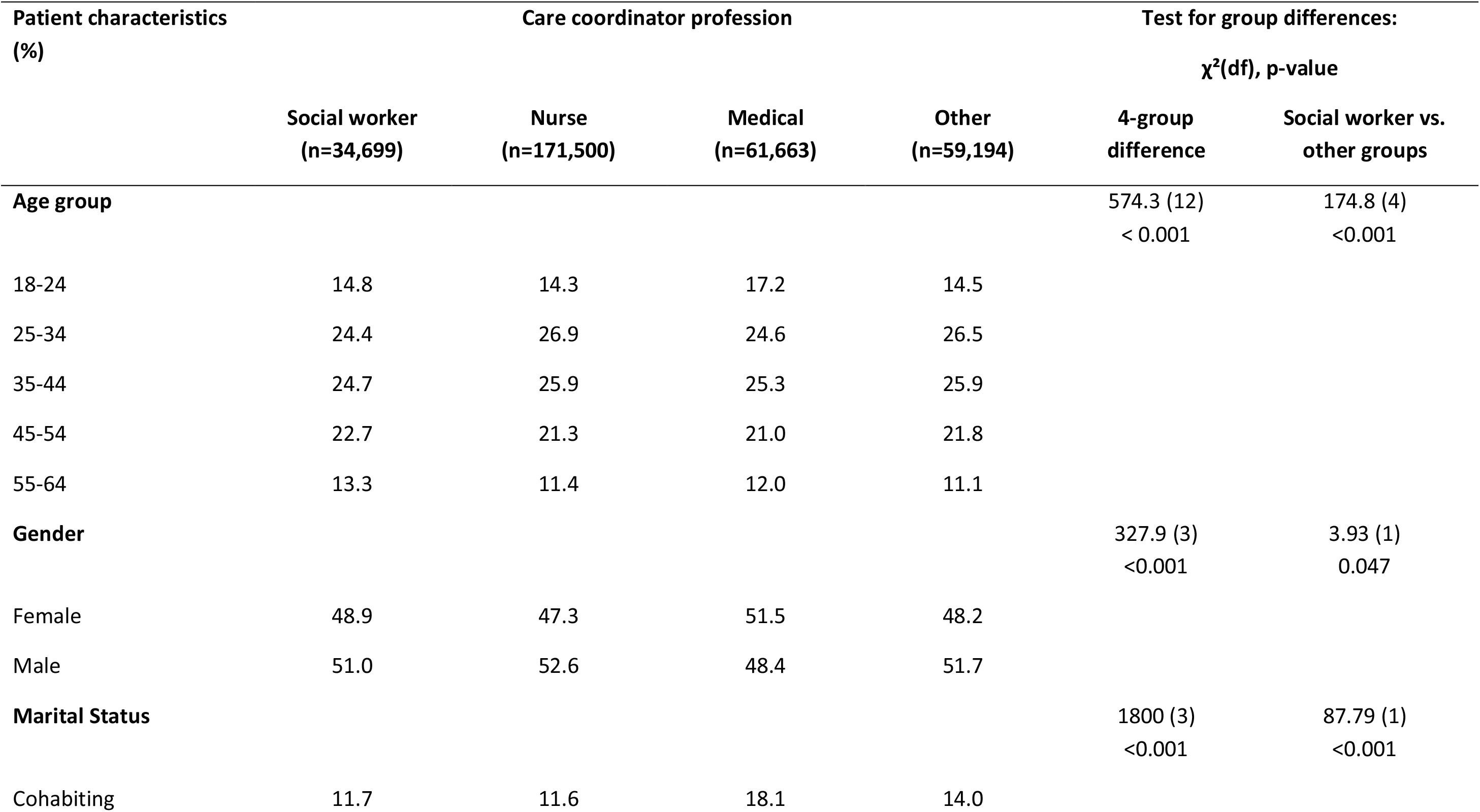
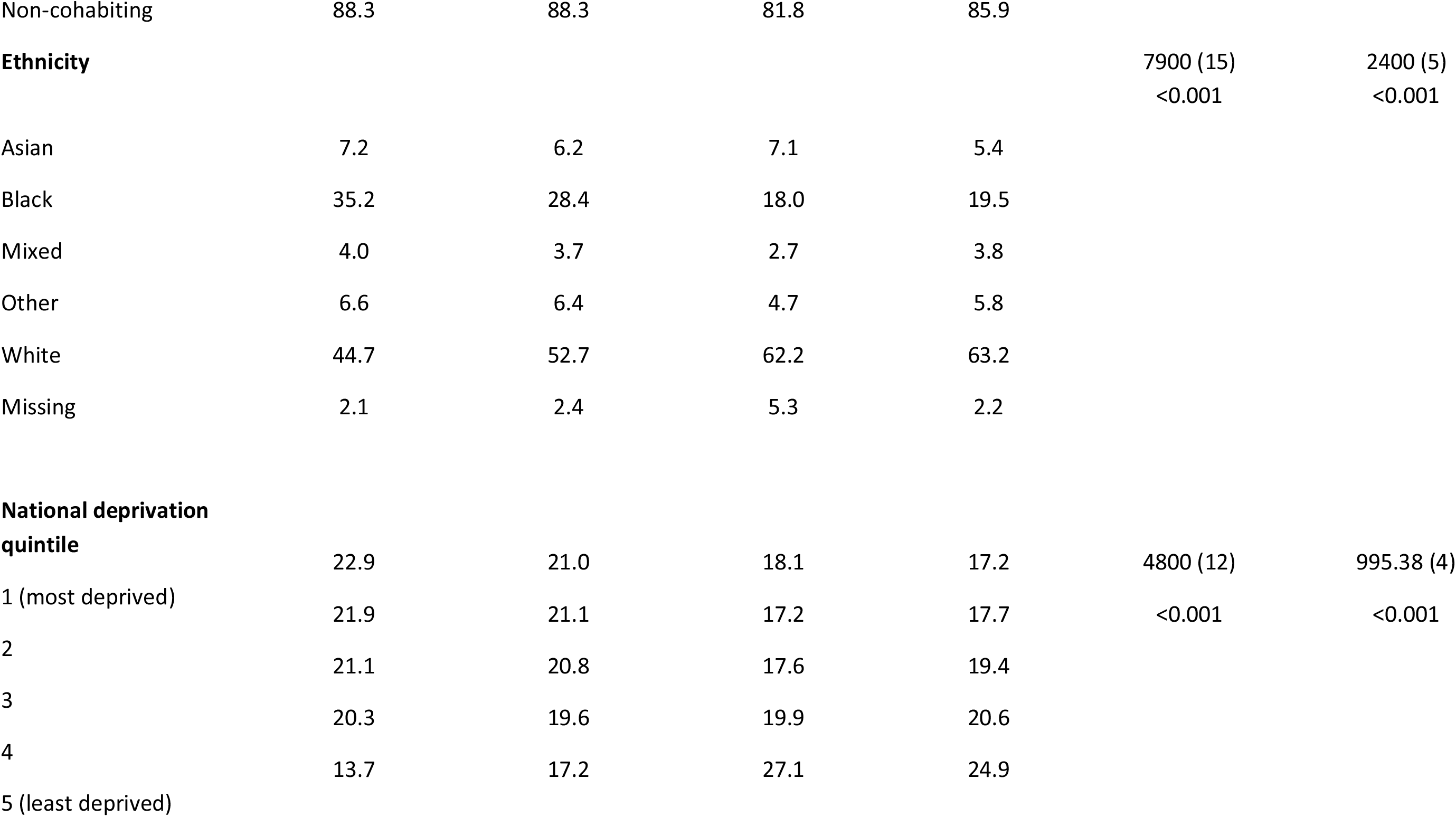
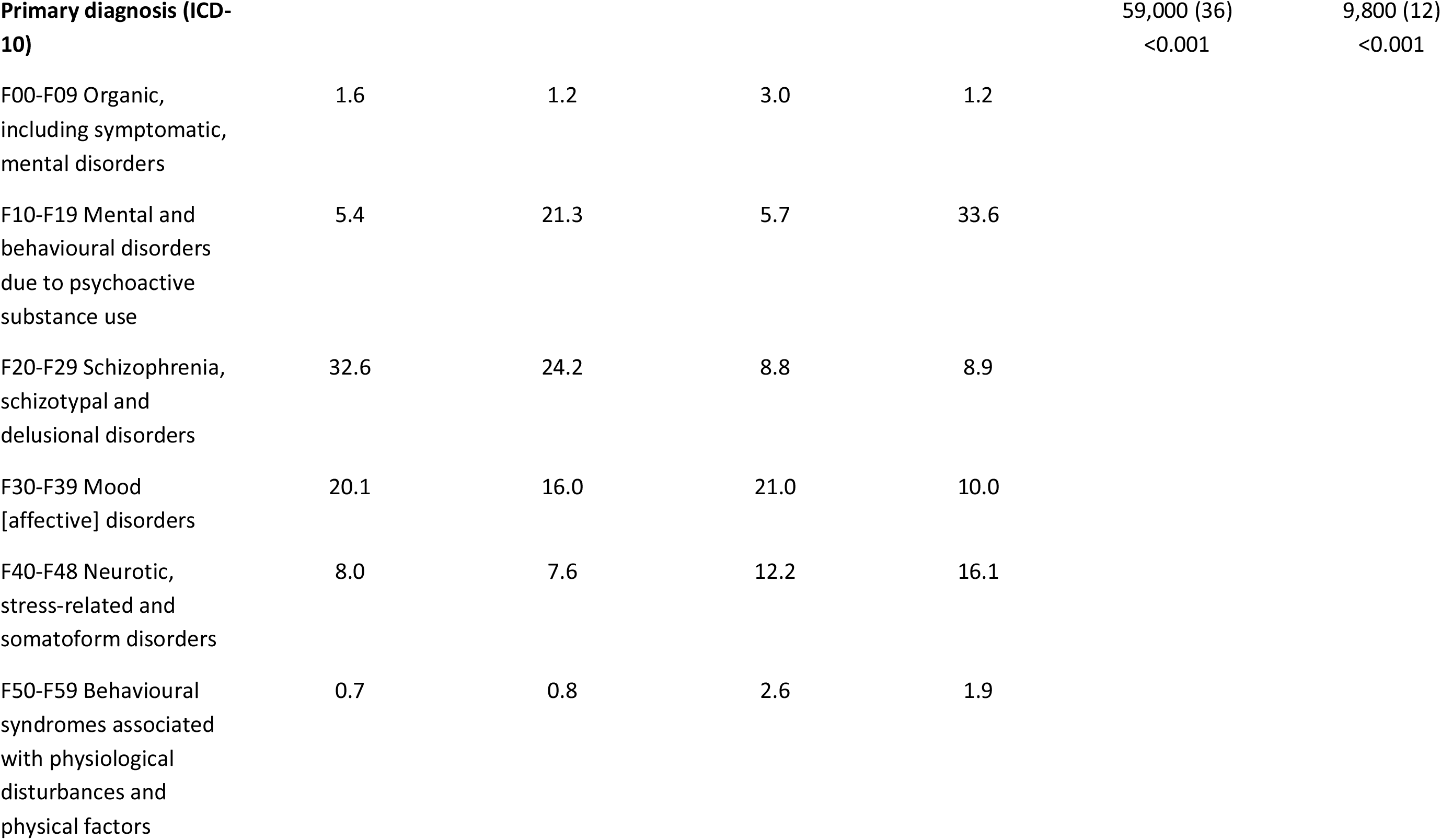
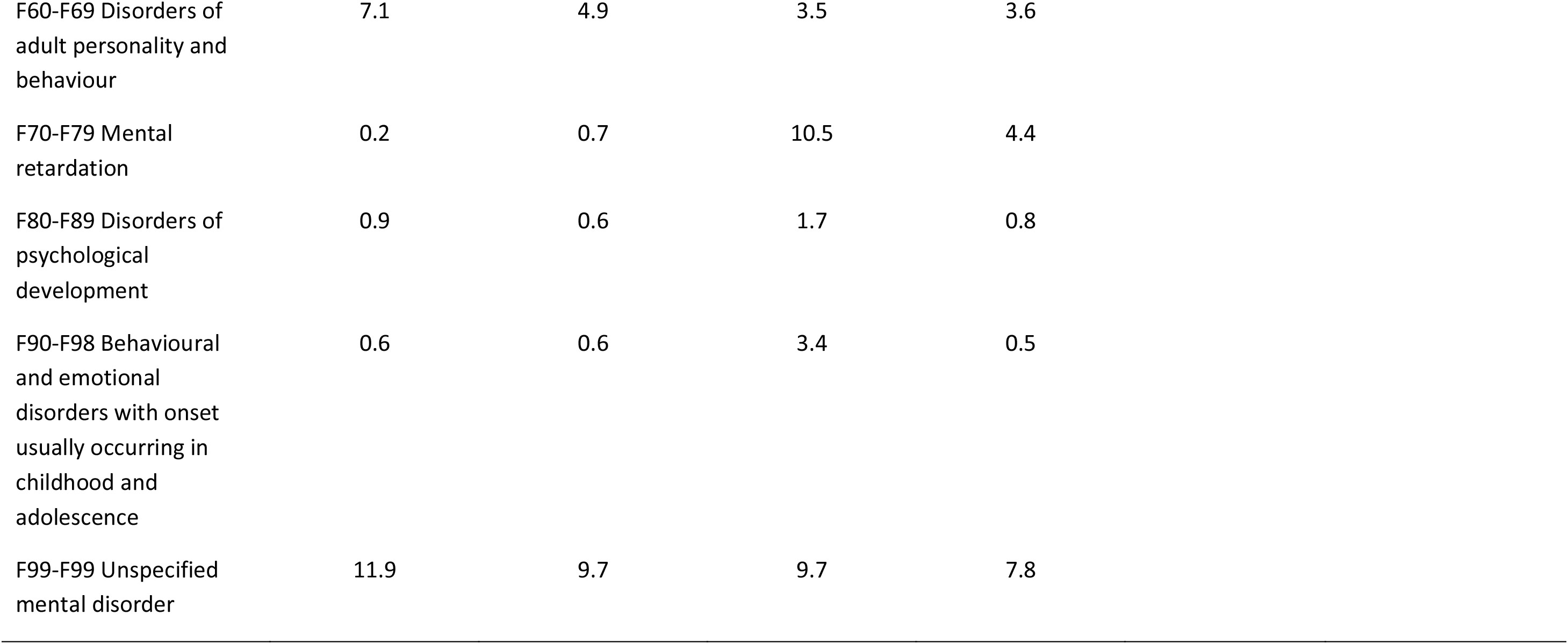
Patient demographic characteristics and primary diagnosis for index care coordination episodes by the professional background of the care coordinator.

Table 2 displays distributions of service contacts prior to the index care coordination episode for the comparison groups. For index episodes with a social worker as care coordinator, mean duration since referral was longer than those for other professionals combined, as was the duration of the care episode itself. Considering the previous 12 months prior to the start date, episodes with a social worker as care coordinator did not differ in number of acute admissions; however, all other measures were more severe – more admissions involving a MHA section, and more days under active mental healthcare, under home treatment team care, and in inpatient care, as well as higher numbers of community and emergency care contacts. Numbers of recorded symptoms in the prior 12 months were also highest for social worker care coordination episodes for all scales, and the most common HoNOS problems were agitated behaviour, hallucinations/delusions, living conditions problems and occupational problems (Table 3).

**Table 2.** Distributions of service contacts prior to the index care coordination episode by the professional background of the care coordinator.

|  | Mean (SD) by care coordinator profession |  |  |  | Test for group difference |  |
| --- | --- | --- | --- | --- | --- | --- |
|  | Social worker<br>(n=34,699) | Nurse<br>(n=171,500) | Medical<br>(n=61,663) | Other<br>(n=59,194) | ANOVA for 4-group<br>difference (df) (F, p-value) | Social worker vs. other<br>groups (df)<br>(t-test, p-value) |
| Duration since referral (years) | 6.55 (6.34) | 5.56 (6.18) | 3.69 (4.65) | 3.83 (5.01) | F (3, 321,516) = 3245.5<br><0.001 | t (321,518) = -52.45<br><0.001 |
| Duration of care coordination episode (days) | 237 (350.3) | 180 (292.7) | 252 (371.8) | 211 (275.6) | F (3, 327,052) = 952.6<br><0.001 | t (327,054) = -19.89<br><0.001 |
| *In the 12 months prior to the index episode* |  |  |  |  |  |  |
| Number of acute admissions | 0.6 (4.1) | 0.6 (2.7) | 0.8 (5.1) | 0.5 (1.8) | F (3, 327,052) = 103.59<br><0.001 | t (327,054) = 0.02<br>0.984 |
| Number of admissions involving MHA section | 0.3 (0.9) | 0.2 (0.7) | 0.0 (0.3) | 0.0 (0.5) | F (3, 327,052) = 2081.0<br><0.001 | t (327,054) = -47.18<br><0.001 |
| Number of days under mental health services | 196.7 (158.9) | 158.6 (158.0) | 162.6 (158.6) | 142.1 (148.0) | F (3, 327,052) = 901.1<br><0.001 | t (327,054) = -45.54<br><0.001 |
| Number of days under home treatment team | 4.4 (14.9) | 2.7 (11.5) | 1.0 (8.9) | 1.3 (8.0) | F (3, 327,052) = 959.9<br><0.001 | t (327,054) = -37.00<br><0.001 |
| Number of days in inpatient care | 19.2 (58.4) | 12.5 (46.9) | 2.4 (20.0) | 5.1 (28.0) | F (3, 327,052) = 1718.1<br><0.001 | t (327,054) = -43.58<br><0.001 |
| Number of face-to-face community contacts | 13.3 (19.7) | 10.3 (18.0) | 4.3 (9.3) | 6.6 (14.0) | F (3, 327,052) = 3236.1<br><0.001 | t (327,054) = -52.99<br><0.001 |
| Number of face-to-face emergency care contacts | 0.6 (2.2) | 0.4 (1.6) | 0.1 (0.7) | 0.2 (1.1) | F (3, 327,052) = 1343.7<br><0.001 | t (327,054) = -36.93<br><0.001 |

**Table 3.**
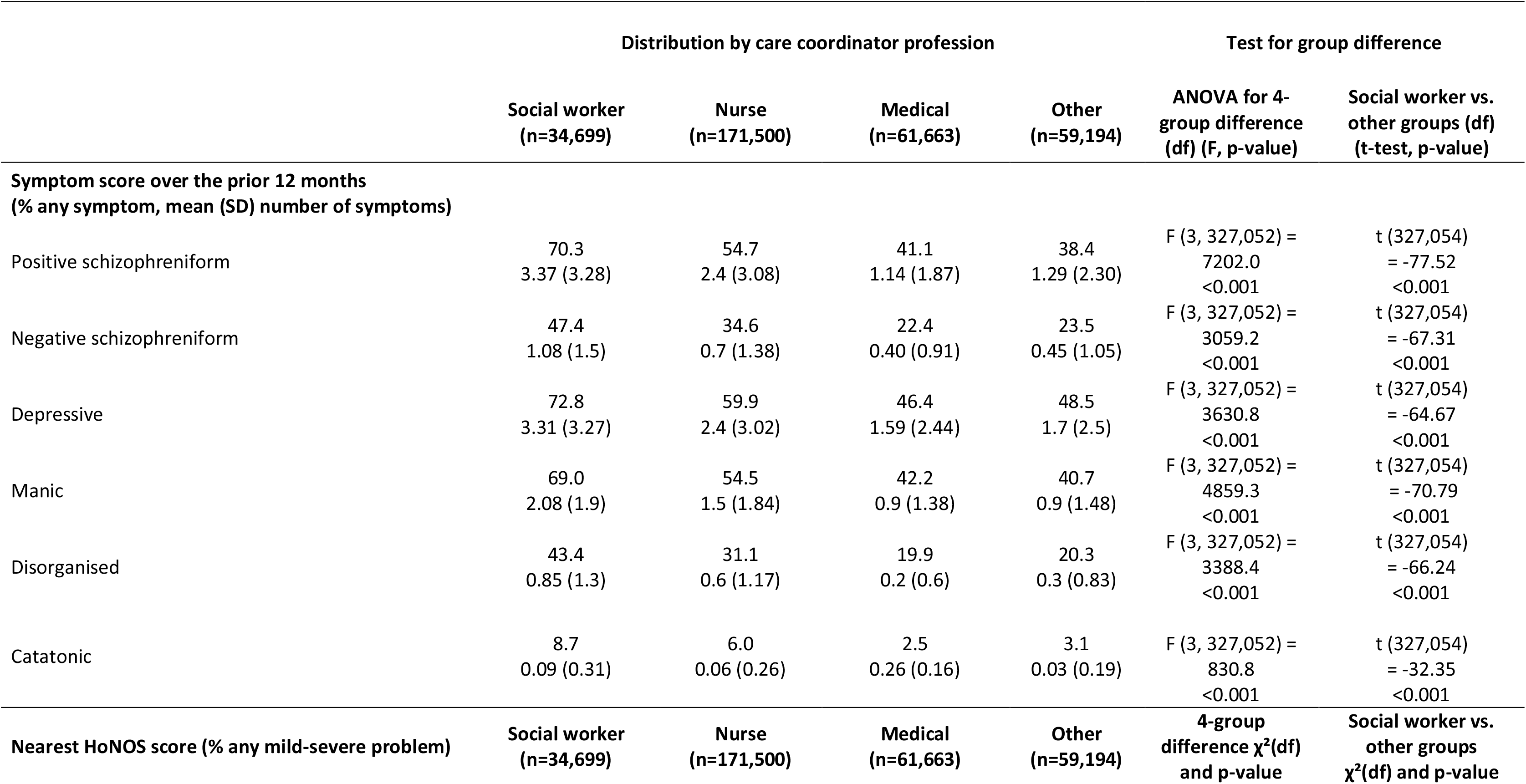
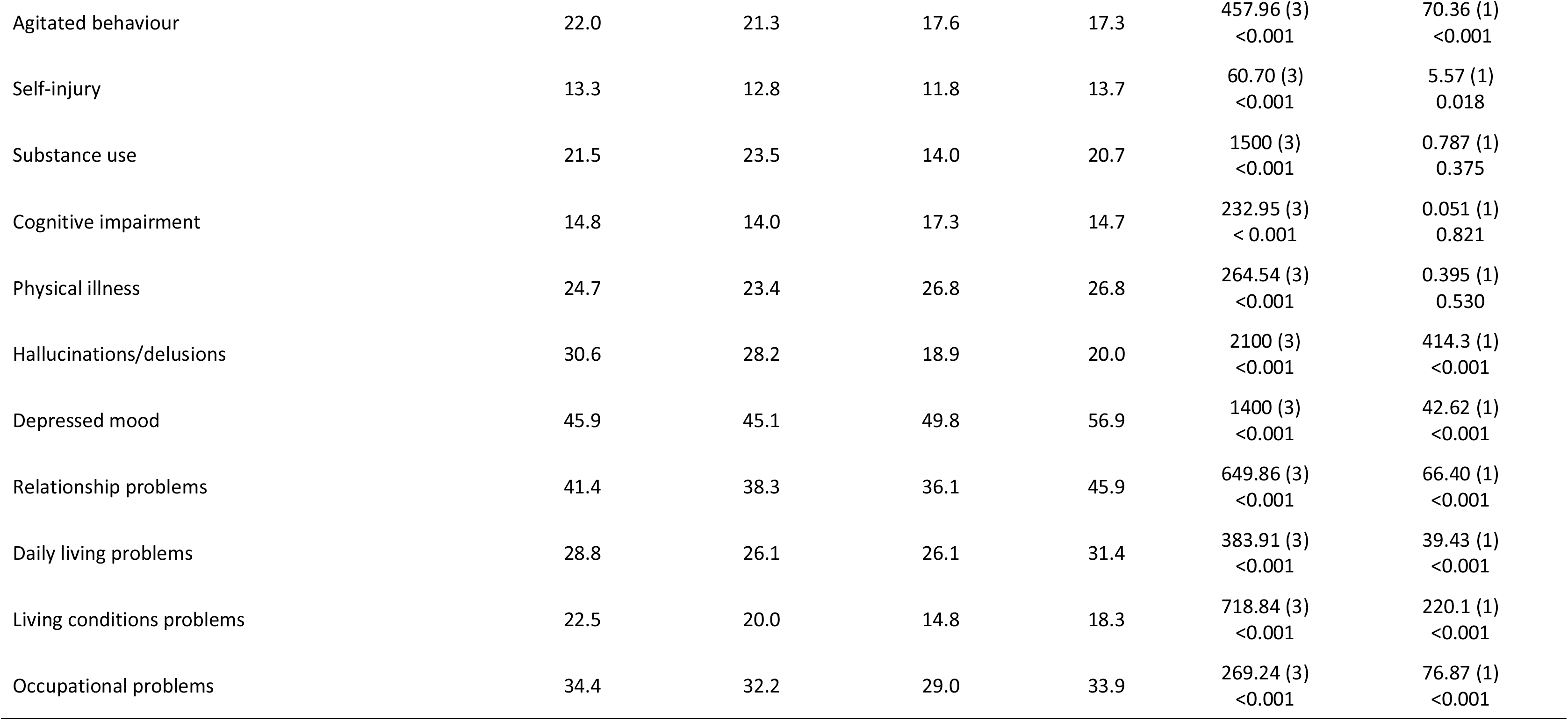
Distributions of clinical characteristics prior to the index care coordination episode by the professional background of the care coordinator.

|  | Distribution by care coordinator profession |  |  |  | Test for group difference |  |
| --- | --- | --- | --- | --- | --- | --- |
|  | Social worker<br>(n=34,699) | Nurse<br>(n=171,500) | Medical<br>(n=61,663) | Other<br>(n=59,194) | ANOVA for 4-group difference<br>(df) (F, p-value) | Social worker vs.<br>other groups (df)<br>(t-test, p-value) |
| **Symptom score over the prior 12 months**<br>**(% any symptom, mean (SD) number of symptoms)** |  |  |  |  |  |  |
| Positive schizophreniform | 70.3<br>3.37 (3.28) | 54.7<br>2.4 (3.08) | 41.1<br>1.14 (1.87) | 38.4<br>1.29 (2.30) | F (3, 327,052) =<br>7202.0<br><0.001 | t (327,054)<br>= -77.52<br><0.001 |
| Negative schizophreniform | 47.4<br>1.08 (1.5) | 34.6<br>0.7 (1.38) | 22.4<br>0.40 (0.91) | 23.5<br>0.45 (1.05) | F (3, 327,052) =<br>3059.2<br><0.001 | t (327,054)<br>= -67.31<br><0.001 |
| Depressive | 72.8<br>3.31 (3.27) | 59.9<br>2.4 (3.02) | 46.4<br>1.59 (2.44) | 48.5<br>1.7 (2.5) | F (3, 327,052) =<br>3630.8<br><0.001 | t (327,054)<br>= -64.67<br><0.001 |
| Manic | 69.0<br>2.08 (1.9) | 54.5<br>1.5 (1.84) | 42.2<br>0.9 (1.38) | 40.7<br>0.9 (1.48) | F (3, 327,052) =<br>4859.3<br><0.001 | t (327,054)<br>= -70.79<br><0.001 |
| Disorganised | 43.4<br>0.85 (1.3) | 31.1<br>0.6 (1.17) | 19.9<br>0.2 (0.6) | 20.3<br>0.3 (0.83) | F (3, 327,052) =<br>3388.4<br><0.001 | t (327,054)<br>= -66.24<br><0.001 |
| Catatonic | 8.7<br>0.09 (0.31) | 6.0<br>0.06 (0.26) | 2.5<br>0.26 (0.16) | 3.1<br>0.03 (0.19) | F (3, 327,052) =<br>830.8<br><0.001 | t (327,054)<br>= -32.35<br><0.001 |
| **Nearest HoNOS score (% any mild-severe problem)** | **Social worker**<br>**(n=34,699)** | **Nurse**<br>**(n=171,500)** | **Medical**<br>**(n=61,663)** | **Other**<br>**(n=59,194)** | **4-group**<br>**difference $\chi^2$(df)**<br>**and p-value** | **Social worker vs.**<br>**other groups**<br>**$\chi^2$(df) and p-value** |
| Agitated behaviour | 22.0 | 21.3 | 17.6 | 17.3 | 457.96 (3)<br><0.001 | 70.36 (1)<br><0.001 |
| Self-injury | 13.3 | 12.8 | 11.8 | 13.7 | 60.70 (3)<br><0.001 | 5.57 (1)<br>0.018 |
| Substance use | 21.5 | 23.5 | 14.0 | 20.7 | 1500 (3)<br><0.001 | 0.787 (1)<br>0.375 |
| Cognitive impairment | 14.8 | 14.0 | 17.3 | 14.7 | 232.95 (3)<br>< 0.001 | 0.051 (1)<br>0.821 |
| Physical illness | 24.7 | 23.4 | 26.8 | 26.8 | 264.54 (3)<br><0.001 | 0.395 (1)<br>0.530 |
| Hallucinations/delusions | 30.6 | 28.2 | 18.9 | 20.0 | 2100 (3)<br><0.001 | 414.3 (1)<br><0.001 |
| Depressed mood | 45.9 | 45.1 | 49.8 | 56.9 | 1400 (3)<br><0.001 | 42.62 (1)<br><0.001 |
| Relationship problems | 41.4 | 38.3 | 36.1 | 45.9 | 649.86 (3)<br><0.001 | 66.40 (1)<br><0.001 |
| Daily living problems | 28.8 | 26.1 | 26.1 | 31.4 | 383.91 (3)<br><0.001 | 39.43 (1)<br><0.001 |
| Living conditions problems | 22.5 | 20.0 | 14.8 | 18.3 | 718.84 (3)<br><0.001 | 220.1 (1)<br><0.001 |
| Occupational problems | 34.4 | 32.2 | 29.0 | 33.9 | 269.24 (3)<br><0.001 | 76.87 (1)<br><0.001 |

Regression model outputs for the different groups of characteristics are displayed in Tables 4-6, all comparing episodes with social worker care coordinators to those with other professionals combined. In fully adjusted models, social worker care coordination remained associated with longer duration since referral, more admissions requiring Mental Health Act, more days under home treatment team, and more face-to-face emergency department contacts and face to face community contacts; number of days under an active mental health service referral, however, days in inpatient care was lower after the final adjustment for symptom scales (Table 4). Recorded symptom scores from the preceding 12 months were higher in all domains (Table 5), and in final adjusted models, social worker care coordination was associated with HoNOS problem-level scores for all subscales apart from substance use, cognitive impairment and physical illness (Table 6).

**Table 4.** Generalised estimating equations (GEE) negative binomial regression for service use prior to the index episode comparing social workers with other care coordinator professionals.

| Service use characteristics in the 12 months prior to the index episode | IRR (95% CI), p-value |  |  |
| --- | --- | --- | --- |
|  | Model 1<sup>a</sup> | Model 2<sup>b</sup> | Model 3<sup>c</sup> |
| Duration since referral (years) | 1.15 (1.13, 1.17)<br><0.001 | 1.14 (1.12, 1.16)<br><0.001 | 1.05 (1.03, 1.07)<br><0.001 |
| Number of acute admissions | 1.02 (0.96, 1.08)<br>0.486 | 1.00 (0.95, 1.07)<br>0.779 | 0.95 (0.90, 1.01)<br>0.169 |
| Number of admissions requiring MHA section | 2.15 (2.07, 2.23)<br><0.001 | 1.92 (1.84, 2.00)<br><0.001 | 1.11 (1.07, 1.15)<br><0.001 |
| Number of days under mental health services | 1.25 (1.24, 1.27)<br><0.001 | 1.19 (1.17, 1.20)<br><0.001 | 0.97 (0.96, 0.99)<br>0.003 |
| Number of days under home treatment team | 1.46 (1.34, 1.59)<br><0.001 | 1.46 (1.34, 1.59)<br><0.001 | 1.25 (1.17, 1.34)<br><0.001 |
| Number of days in inpatient care | 2.16 (2.06, 2.27)<br><0.001 | 2.00 (1.90, 2.11)<br><0.001 | 1.04 (0.95, 1.14)<br>0.317 |
| Number of face-to-face community contacts | 1.59 (1.56, 1.62)<br><0.001 | 1.52 (1.48, 1.55)<br><0.001 | 1.03 (1.01, 1.06)<br>0.005 |
| Number of face-to-face emergency contacts | 1.36 (1.26, 1.47)<br><0.001 | 1.35 (1.26, 1.45)<br><0.001 | 1.10 (1.07, 1.14)<br><0.001 |

<sup>a</sup> Model 1: Unadjusted
<sup>b</sup> Model 2: Adjusted for gender, ethnicity, marital status, age and IMD
<sup>c</sup> Model 3: Adjusted for model 2 + primary diagnosis and symptoms score one year prior the recorded care coordination

**Table 5.** Generalised estimating equations (GEE) negative binomial regression for symptom score prior to the index episode comparing social workers with other care coordinator professionals.

| Symptoms score in the 12 months prior to the index episode | IRR (95% CI), p-value |  |  |
| --- | --- | --- | --- |
|  | Model 1<sup>a</sup> | Model 2<sup>b</sup> | Model 3<sup>c</sup> |
| Positive schizophreniform | 1.75 (1.73, 1.78)<br><0.001 | 1.64 (1.62, 1.67)<br><0.001 | 1.13 (1.10, 1.15)<br><0.001 |
| Negative schizophreniform | 1.18 (1.13, 1.23)<br><0.001 | 1.19 (1.15, 1.23)<br><0.001 | 1.15 (1.12, 1.18)<br><0.001 |
| Depressive | 1.17 (1.14, 1.19)<br><0.001 | 1.17 (1.15, 1.20)<br><0.001 | 1.15 (1.13, 1.17)<br><0.001 |
| Manic | 1.62 (1.60, 1.64)<br><0.001 | 1.13 (1.11, 1.16)<br><0.001 | 1.14 (1.12, 1.16)<br><0.001 |
| Disorganised | 1.79 (1.75, 1.83)<br><0.001 | 1.17 (1.12, 1.22)<br><0.001 | 1.15 (1.12, 1.18)<br><0.001 |
| Catatonic | 1.26 (1.14, 1.40)<br><0.001 | 1.26 (1.16, 1.37)<br><0.001 | 1.18 (1.11, 1.25)<br><0.001 |

<sup>a</sup> Model 1: Unadjusted
<sup>b</sup> Model 2: Adjusted for gender, ethnicity, marital status, age and IMD
<sup>c</sup> Model 3: Adjusted for model 2 + primary diagnosis and service use variables (acute admissions, days under home treatment team and face-to-face community contacts) one year prior the recorded care coordination

**Table 6.** Logistic regression models for the nearest HoNOS score prior to the index episode comparing social workers with other care coordinator professionals.

| Nearest HoNOS score in the 12 months prior to the index episode | Odds ratio (95% CI), p-value |  |  |
| --- | --- | --- | --- |
|  | Model 1<sup>a</sup> | Model 2<sup>b</sup> | Model 3<sup>c</sup> |
| Agitated behaviour | 1.13 (1.09, 1.16) <0.001 | 1.11 (1.07, 1.14) <0.001 | 1.07 (1.04, 1.10) <0.001 |
| Self-injury | 1.04 (1.01, 1.08) 0.018 | 1.09 (1.05, 1.14) <0.001 | 1.13 (1.09, 1.18) <0.001 |
| Substance use | 1.01 (0.98, 1.04) 0.375 | 0.99 (0.96, 1.02) 0.788 | 1.03 (1.00, 1.07) 0.028 |
| Cognitive impairment | 1.01 (0.98, 1.04) 0.375 | 0.99 (0.96, 1.02) 0.788 | 1.03 (1.00, 1.07) 0.028 |
| Physical illness | 1.00 (0.98, 1.03) 0.530 | 0.97 (0.94, 1.00) 0.113 | 1.03 (1.00, 1.06) 0.045 |
| Hallucinations/delusions | 1.31 (1.27, 1.34) <0.001 | 1.21 (1.18, 1.25) <0.001 | 1.09 (1.06, 1.12) <0.001 |
| Depressed mood | 0.92 (0.90, 0.94) <0.001 | 0.98 (0.95, 1.00) 0.169 | 1.06 (1.03, 1.09) <0.001 |
| Relationship problems | 1.10 (1.07, 1.13) <0.001 | 1.09 (1.07, 1.12) <0.001 | 1.09 (1.06, 1.12) <0.001 |
| Daily living problems | 1.08 (1.06, 1.11) <0.001 | 1.05 (1.02, 1.08) <0.001 | 1.08 (1.05, 1.11) <0.001 |
| Living conditions problems | 1.24 (1.21, 1.28) <0.001 | 1.18 (1.15, 1.22) <0.001 | 1.17 (1.14, 1.21) <0.001 |
| Occupational problems | 1.11 (1.09, 1.14) <0.001 | 1.07 (1.04, 1.10) <0.001 | 1.07 (1.04, 1.09) <0.001 |

<sup>a</sup>Model 1: Unadjusted<sup>b</sup>Model 2: Adjusted for gender, ethnicity, marital status, age and IMD<sup>c</sup>Model 3: Adjusted for model 2 + primary diagnosis and symptoms score one year prior the recorded care coordination

## Discussion

We investigated differences in characteristics by care coordinator profession in 327,056 recorded care coordination episodes for 132,553 patients aged 18-65 years, with a particular focus on social workers as a professional group. From the large sample, 10.6% of the recorded episodes had a social worker as a care coordinator and a range of mutually independent differences in characteristics were observed, tending towards more complex and severe needs as measured prior service use and recorded symptom profile and by the closest recorded HoNOS problems scales.

Care coordination is a fundamental element of mental healthcare and integrated services, providing comprehensive care that involves medical support and social care, especially for people with complex needs.^7,15^ Support for people with mental disorders can be managed and organised by members of multidisciplinary teams, e.g., social workers or nurses, across different settings.^3,7^ However, a study by Hannigan et al. (2018) conducted interviews on daily tasks with 28 care coordinators from diverse professional backgrounds across six community mental health services in England and Wales and reported that, in practice, care coordinator allocations may not be based on people’s needs, and services may be delivered by system pressure, caseload, and team structures.^16^

Lately, there has been greater attention to the importance of social needs in healthcare, particularly mental healthcare.^7,15,17^ Social workers can play a vital role in mental healthcare by coordinating care and providing psychosocial support that addresses social determinants.^3,17^ However, few quantitative studies have explored care coordination using routine mental health data. One large-scale study investigating patient-sharing networks between healthcare providers (hospitals, GP, psychiatrists, nurses), considered the care coordination using French national administrative data extracted for people aged 20-56 diagnosis with a severe mental health disorder who had been hospitalised at least once; data were obtained one year after the end of the hospitalisation to investigate impact on care quality.^18^ They found that although there were varied types of patient-sharing networks, the structure alone did not associate with better outcomes.^18^ However, the data were collected before national mental health policy reforms (2016), when the system moved to stronger integration of health and social care; therefore, the study did not capture social care.^18^ In an English national survey of 122 non-statutory organisations providing diverse services, 17 services were selected for both rural and urban areas that completed at least 4 of 8 care coordination tasks.^19^ The study found that service providers were most of the time in direct contact with service users/carers, and the mean weekly cost per episode was around 69.7 GBP for the full sample.^19^ The study also noted that non-statutory care coordinators overlapped with social work but without statutory authority. However, the study mainly investigated the care coordination tasks, time, and costs; also, 49% of the supported group were people with dementia.^19^

A key finding of our analysis is that, compared to other professionals, social worker care coordination tended to occur later in the presentation, commencing at a longer mean duration since the original referral. These findings are consistent with social worker reports that many patients are not receiving the help and support they need at the right time.^17,20^ Furthermore, Walton et al. (2023) conducted a cross-sectional national survey to examine the care coordination experiences of 1,457 people with rare diseases, carers, and professionals in the UK, finding that service users and carers reported limited access to care coordination, while professionals reported that their patients had better access to care coordination.^21^

Several measures of prior service use were higher for social worker care coordination episodes: specifically, the number of days of admissions involving a compulsory order or under a home treatment team, as well as higher face-to-face emergency care contacts, higher recorded symptoms and higher likelihood of problem scores on most HoNOS scales. Our results were consistent with the study by Moore et al. (2016), which investigated the role of social workers in Mental Health Services and Care Coordination in a large urban Trauma Centre Emergency Department in the USA; in a retrospective review of 49,354 visits over 15 months, this study found that social workers, supporting 38% patients, were working with more complex cases medically and socially.^22^ Moreover, in a previous analysis of social worker contacts from 2008 to 2024 in 260,844 patients aged 16 and older, we found that 20% had at least one social work contact and the majority were in people diagnosed with schizophrenia (39.2%) and living in the most deprived areas.^14^ Our results therefore suggest that complex cases and more severe symptoms predict social worker care coordination, and that there may be a more purposeful allocation of care coordination roles than has been reported elsewhere.^14^ However, as noted earlier, there are still limited quantitative investigations of care coordinator roles generally, including social worker roles specifically, using large-scale mental health data, and findings require wider evaluation.

## Strengths and limitation

This study has several strengths, including the use of a large sample from routine mental healthcare over a long period, evaluating a range of professional care coordination roles and from a highly socially and culturally diverse catchment. As well as service data, we were able to incorporate an evaluation of symptom profiles utilising natural language processing and, we believe, the first study to do so at this scale. A key limitation, as mentioned, is the single catchment and provider, so that replication is needed in other settings, although we believe that component services have substantial diversity, even more so at the level of individual care coordinator, so we would not expect a substantial top-down influence from SLaM as the single Trust provider. Comparisons relied on the assigning of a profession to each care coordinator, which will not have been totally accurate; however, misclassification would not obviously have been influenced by any of the comparison variables and hence would have obscured rather than exaggerated observed associations.

Considering confounding factors, a range of sociodemographic and clinical variables were extracted and utilised; however, there are important characteristics and features that were not included, including levels of social support and individual-level (rather than area-level) poverty. Clinical covariates were additionally limited to the 12-month period prior to the episode of care, hence did not capture longer-term history. Finally, we did not attempt to adjust for individual-level teams, so it is possible that at least part of the comparison is between caseloads of teams with/without social worker care coordinators rather than caseloads of care coordinators themselves. Findings are derived from observational data hence causal associations cannot be assumed.

## Conclusion

This study compared care coordination episodes according to the profession of the care coordinator, focusing on social workers, using large-scale data from routine mental health services in South London. The study found that patients with social workers as care coordinators, compared to other professional groups, have more complex past clinical needs and severe symptoms, who are being seen later in the course of mental healthcare. This study contributes to improving our understanding of mental health care coordination in real-world practice; however, the process by which care coordinators are assigned remains unclear, and further research is needed to better understand this. Large, granular health records datasets could help further clarify such processes, although need to be complemented by in-depth qualitative research.

## Data Availability

No data are available. Data may be obtained from a third party and are not publicly available. Access to deidentified data can be applied for via the NIHR Maudsley Biomedical Research Centre at the South London and Maudsley NHS Foundation Trust, upon reasonable request. Requests for data will be considered on a case-by-case basis, given the sensitive nature of the data, and access will only be granted if approval is given by the Work and Health Screening Panel and other governance requirements are fulfilled. For more information, please contact

https://projects.slam.nhs.uk/about/

## Statements and Declarations

## Acknowledgements

We would like to thank the Saudi Ministry of Education for its generous financial support through the Cultural Bureau of the Royal Embassy of Saudi Arabia.

## Author Contributions

The study was conceived by NA and RS. The data were extracted by MB. Analyses were carried out by NA and GP. The manuscript was written by NA under RS and GP supervision and finalised with input from all authors.

## Ethics approval

Oxford Research Ethics Committee C, reference 23/SC/0257.

